# Effect of Pranayama on Perceived Stress, Well Being and Quality of Life of Frontline Healthcare Professionals on Covid-19 Duty: A Quasi-Randomized Clinical Trial

**DOI:** 10.1101/2022.08.25.22279201

**Authors:** Rakesh Sarwal, Rajinder K. Dhamija, Khushbu Jain, Ishwar V. Basavaraddi

## Abstract

**Background:** The COVID-19 pandemic has brought unparalleled challenges for health systems worldwide, the impact of which has also been borne by the Healthcare Professionals (HCPs). Numerous studies have revealed the positive effects of Pranayama and Meditation on mental health. The effect of Pranayama in improving mental health of frontline HCP exposed to Covid-19 patients has not been studied.

**Aim & Objective:** This quasi-randomized clinical trial was done to study the effect of especially designed Pranayama protocol on perceived stress, wellbeing and quality of life of frontline health care professionals who were exposed to COVID-19 patients in hospital settings.

**Methodology:** This study was done with 280 frontline healthcare professionals (HCP) assigned duties with COVID-19 patients during September-November, 2020 in 5 government hospitals and COVID-19 quarantine/isolation centres in New Delhi, India. The HCPs were first assessed for COVID-19 infection in the past using antibody test, and only those found negative were recruited. The enrolled respondents were randomly assigned to two arms – an intervention arm where there were to practice 28-day Pranayama module (morning and evening sessions) under supervision of a trainer, and a Control arm where the HCPs continued routine physical activity (walking, jogging etc.). Baseline and end-line (total: 250 HCPs) Psychological parameters of Perceived Stress, Well Being and Quality of Life were collected through self-reported questionnaires.

**Results:** The intervention (HCPs: 123) and control (HCPs: 127) groups (Total: 250) were comparable in their demographic profile and baseline characteristics. Intervention with Pranayama module led to a significant reduction (Mean diff: -2.46; P-value: 0.028) in perceived stress score in the intervention group compared to the control group. The wellbeing index in Interventional group intervention showed a non-significant increase. The WHO Quality-of-life score increased in the intervention group as compared to the controls (mean difference 2.78, p-value: 0.17). Of its four components, the one for Psychological domain increased significantly (mean diff: 1.52, P-value: 0.019), while those for Physical domain and Environmental domains increased (mean diff: 0.64, P-value: 0.29 and mean diff: 0.68, p-value: 0.48) though not statistical significantly.

**Cconclusion:** The intervention of twice daily **p**ractice of the Pranayama module for 28 days in frontline HCPs performing COVID-19 duties had a noteworthy effect in lowering Perceived Stress, improving perceived Quality of life, especially its Psychological domains as measured through standardized questionnaires.

**CTRI Number:** **CTRI/2020/07/026667**

## INTRODUCTION

Like other pandemics and disease outbreaks, COVID-19 has created immense psychosocial disturbances for the healthcare workers. During a pandemic outbreak of a global scale, frontline HCPs, who are in direct contact with the patients are exposed to the highest levels of risk (WHO2003). Nurses and other allied healthcare workers are particularly vulnerable to many job-related hazards, and thereby undergo a considerable amount of emotional pressure (Wheeler 1997).

The COVID-19 has had significant negative impacts on health-care workers’ psychological health, triggering anxiety, depression, and sleep disturbance (Chirico Fet.al.2021). Thus, frontline health-care workers need psychological support, through means as occupational health surveillance programs that train and educate health-care workers to raise their capacity to address the risk of infectious disease and associated psychological distress (Chirico Fet.al.2021). Various therapies like TCM (Traditionally Chinese medicine) (Ren JL et al., 2020). Ayurveda has also been used for the management of Covid-19 related stress (Maurya VK.et. al. 2020)

Yoga based intervention are known to lower stress and improve quality of life in many conditions. A 12-weeks of yoga-based lifestyle intervention in the parents of retinoblastoma patients lead to a significant improvement in psychological stress and overall quality of life (Bisht S et.al.2019). There was a significant improvement in all the domains (physical health, psychological health, social relationships, and environment) of World Health Organization Quality of Life- Brief (WHOQOL-BREF) within one month of yoga therapy (Bisht S., 2019).

Numerous studies have examined ways of reducing stress and burnout in HCPs, through programs that incorporate mind-body practices, honing cognitive skills, community building, and connecting with meaning and purpose in work (Shanafelt TD 2017; West CP 2016; Lee HF 2016; Scheepers RA 2020). There is strong evidence on the positive effects of Yoga practice on stress management among HCPs (Bremner JD et al.202; Chu IH et al.2017). Many recent studies have demonstrated the positive impact of various yoga-based interventions in different HCPs population (Cocchiara, RA 2019; La Torre G 2020; Ofei-DodooS et. al. 2020; Rostami, K et.al.2019). A recent study on 40 health care workers of two hospitals in Rome showed significant decrease in their stress and anxiety scores and increase in quality-of-life assessment scores after 4 weeks of yoga and mindfulness course. (Torre GL et al. 2020). A pilot study conducted in United States demonstrated the positive effect of group mindfulness-based yoga activities on personal accomplishment, depression, anxiety, stress, perceived resilience, and compassion in HCPs (Ofei-Dodoo, Set. al.2020). In another pilot study on ICU nurses, those who received yoga instructions did better on the measures of quality of life after the intervention (Rostami, K et.al.2019). Another study found yoga to be more effective than cognitive training programs in achieving mental well-being and a reduction of stress-related consequences (Riley K.E. et.al. 2017). Yet another study found a significant (*P*<0.001) decrease in anxiety and depression scores, as well as improvement I quality-of-life among the study participants in the yoga group as compared with the control group (Umadevi P., 2013).

Hence, we hypothesised that Yoga can be an effective practice to reduce mental stress in frontline HCPs exposed to COVID-19. The present study was therefore carried out to evaluate the effect of pranayama on Perceived Stress, Well Being and Quality of Life of frontline Health-care Professionals exposed to COVID-19 as a part of their professional duties.

## METHODOLOGY

### Ethical Considerations

After obtaining approval of the Institutional Ethics Committee at Morarji Desai National Institute of Yoga (MDNIY), New Delhi, this study was registered on the Clinical Trials Registry of India (CTRI No. **CTRI/2020/07/026667)**. The government of National Capital Territory of Delhi accorded permission for conduct of the study during September-November 2020 in five hospitals dedicated for Covid19 patients. Informed consent was obtained from the study participants at enrollment. One Yoga instructor was assigned to each of the five identified hospitals in Delhi to train, guide, supervise the participants in the intervention of twice daily practice of Pranayama and also to collect the data. Due to Covid-19 protocols and to avoid risk of exposure, the training was conducted virtually through Video-Conference. The HCPs in the control group were advised general fitness practices (like walking, jogging, running), which they were allowed to practice unsupervised.

### Sample Size

Sample size was estimated on the assumption of a prevalence of COVID-19 infection of 10% in the population at that time, and expectation that our treatment group will have a 90% lower prevalence i.e., 1%. For a power of 80%, alpha of 0.05, 95% confidence interval, the sample size estimate was 121 in each arm (Table 3).

### Study Sample

Inclusion criterion in our study was being a HCPs (age range: 19-65 years, any gender) assigned to and performing COVID-19 duty (duty cards were taken as proof) and free of recent or past COVID-19 (negative antibody test). 288 HCPs were registered for the study.

Out of these, 8 were found positive on rapid Antibody tests at baseline, indicating previous infection of COVID-19, and were therefore excluded from the study. The remaining 280 participants were enrolled for the study. All the HCPs were unvaccinated (Vaccine was not invented)

### Randomization

Medical Officers in-charge of the hospitals/COVID-19 care units were authorized to enroll study participants, but had no other role in the trial. Eligible HCPs were quasi-randomized on enrolment in the trial through alternate allocation (ratio 1:1) into two groups. Subsequently, the two groups were designated as ‘Intervention group’ and the ‘Control group’ by the coordinator of the study, who was blinded to the initial allocation.

### Masking

It is difficult to assess yoga/pranayama practices in double blind trials because the intervention requires active involvement of the participants and hence their identities become known after allocation.

### Study groups

Of the 280 enrolled participants, 17 (Intervention - 15, Control - 2) did not adhere to the 80% attendance requirement, and 13 (Intervention - 2, Control - 11) did not give their post-intervention data, and thus 30 study participants (Intervention - 17, Control - 13) were excluded. Thus, 250 participants completed the study which included 123 in the intervention group (administered Pranayama protocols) and 127 in the control group. Profession wise break up of 250 participants was: Resident Doctors-29, Nursing staff-63, Caretakers / House-keeping staff-82, Lab technicians-22, CMOs/Medical Superintendent -20, Pharmacist-15, Yoga Instructors-4, Hospital Administration staff like Accountants, Multi- Tasking Staff, Data Entry Operator, Receptionists -15.

### Study Parameters and Tools

Basic demographic details, medical history and dietary habits were collected at the beginning of the study. Baseline and end-line data was collected from all the study participants through predesigned, validated questionnaire (which were made available in both Hindi and English Languages) on their Perceived Stress, Well Being and Quality of Life. Antibody test for COVID-19 was done on all the study participants at baseline and at the end of the study.

### Perceived Stress Scale (PSS)

PSS Scale developed by Sheldon Cohen and Williamson (1988) is used to assess the level of stress. The scale compresses of 10 items designed to capture stress through 5 points, with the answer scale ranging from 0 (did not apply to me at all) to 4 (apply to me very much or most of the time). Therefore, the possible scores range from 0 to 40 The scale has a high internal consistency and reliability (alpha=.78) and a moderate concurrent criterion validity with the amount of stress experienced during an average week (r= .39, p<.001) (Sharon et.al. 2019).

### WHO Well-being Index

WHO-5 Well-Being Index introduced by the Regional Office of WHO for Europe (1988) was used to assess the status of mental health. The scale compresses of 5 items, with 6 points answer ranging from 0 (did not apply to me at all) to 5 (apply to me very much or most of the time). Therefore, the possible scores range from 0 to 25. Internal consistency of the WHO-5 scale is good (Cronbach’s alpha = 0.858) (Reza et. al. 2019)

### Quality of Life (WHO-QOL Brief)

Quality of Life (WHO-QOL) Scale developed by the WHO-QOL group (1995) comprises of 26 items, further categorized into four domains of Physical, Psychological, Social and Environmental. It is also a 5-point scale with responses ranging from 1 (did not apply to me at all) to 5 (apply to me very much or most of the time) and vice-versa in few questions (Question 3, 4 and 5). Therefore, the possible scores range from 38 to 118. Internal consistency and reliability for the overall WHOQOL score is high (Cronbach’s alpha= 0.896) (Jahanlou et. al. 2011). The WHOQOL-BREF consists of four domains: physical health (7 items: minmax scores: 15-27), psychological health (6 items: minmax scores: 10-26), social relationships (3 items; minmax scores: 3-15), and environmental health (8 items; minmax scores: 8-40); it also contains QOL and general health items. The domain scores are not averages, they are the sum total score for each question within the domain. The psychological domain with 6 questions/covers aspects like bodily image and appearance, Negative feelings, Positive feelings, Self-esteem Spirituality / Religion / Personal beliefs Thinking, learning, memory and concentration of an individual, therefore covers mental health of a person. The physical domain of WHO QOL with its 7 questions covers activities of daily living, dependence on medicinal substances and medical aids, Energy and fatigue, Mobility, Pain &discomfort Sleep and rest, work Capacity was assessed which was the important criteria during COVID-19 pandemic. Environmental domain with 8 questions/items assesses the financial resources, Freedom, physical safety and security Health and social care, accessibility and quality Home environment Opportunities for acquiring new information and skills Participation in and opportunities for recreation / leisure activities Physical environment (pollution / noise / traffic / climate) and Transport.

Questionnaires has been provided as supplementary data (Annexure-I)

### The Intervention: Administration of Pranayama Protocols

Two Pranayama modules lasting 30 minutes for the morning (Table-1) and 15 minutes for the evening sessions (Table-2), were developed by the Principal Investigator, Participants in the intervention group were trained and guided by Yoga Instructors through Video-Conference to practice Pranayama twice a day for 28 days during the study period of September-November 2020. A daily attendance record was maintained. All the participants were closely monitored by the Yoga Instructors for confirmation to the protocol. The control group was advised general fitness practices (like walking, jogging, running).

**TABLE-1:** Pranayama (Breathing) Protocol for the Morning Session (30 Minutes)

| S. No | Practices | Name of Practice | Rounds | Duration<br>(in Minutes) |
| --- | --- | --- | --- | --- |
| 1 | Preparatory Practices for Pranayama<br>(9 minutes) | Prayer | 3 deep breathing or prayer of individual | 1 |
| 2 |  | Vaata-Neti | 3 rounds* (30 secs/round) | 2.5 |
| 3 |  | Kapalabhati | 3 rounds* (30 secs/round) | 2.5 |
| 4 |  | Deep Breathing | 10 rounds | 3 |
| 5 | Pranayama Practices<br>(16 minutes) | Nadi-shodhana | 10 rounds | 8 |
| 6 |  | Ujjaayee | 10 rounds | 4 |
| 7 |  | Bhramari | 10 rounds | 4 |
| 9 | Meditation<br>(5 minutes) | Dhyana | Awareness of breathing, thoughts and emotions | 5 |
|  | Total Duration |  |  | 30 |

Note: \* (2 &3) Each practice of vaataneti/kapalbhati has suitable rest/ gap time of 20 seconds to become normal and to experience the impact of the practice.
This Pranayama protocol was administered with increased intensity gradually to achieve 6:3:6:3 ratio (6 inhalations: 3 retentions: 6 exhalations: 3 retentions)
Day 1: subjects were practiced Inhalation (4 seconds): Exhalation (4 seconds)
Day 2: I: E ratio (5 seconds :5 seconds)
Day 3: I: E ratio (6 seconds :6 seconds)
Day 4 & 5: I: RI:E ratio (6 seconds :3 seconds :6 seconds)
Day 6 & 7: I: RI: E: RO ratio (6 seconds :3 seconds :6 seconds :3 seconds)
8 th day onwards they continuously practiced with 6 seconds of inhalations: 3 seconds of retentions: 6 seconds of exhalations: 3 seconds of retention ratios

**TABLE-2:** Pranyama (Breathing) Protocol for the Evening Session (15 Minutes)

| S. No | Name of Practice | Rounds | Duration<br>(in Minutes) |
| --- | --- | --- | --- |
| 1 | Shavasana (Corpse Pose) (with palm upwards)<br>Prithabhami Tadasana |  | 1 |
| 2 | Abdominal Breathing | 15 Rounds* | 3 |
| 3 | Thoracic Breathing | 15 Rounds* | 3 |
| 4 | Clavicular Breathing | 15 Rounds* | 3 |
| 5 | Deep Breathing (lying down position) | 15 Rounds* | 3 |
| 6 | Relaxation in Shavasana with awareness on Abdominal breathing |  | 2 |
| Total Duration |  |  | 15 |

Note: Each breathing comprises 6 seconds inhalation and 6 seconds exhalation.
\*Initially the practice of all 4 breathing practices started with 10 rounds each and gradually over a period of 5-7 days increased to 15 rounds each.
There is no retention of breathing in the evening session. For the breathing practices in the evening yoga sessions were designed as follows:
Day 1 & 2: subjects were practiced Inhalation (4 seconds): Exhalation ratio (4 seconds)
Day 3 & 4: Inhalation (5 seconds): Exhalation ratio (5 seconds)
Day 5 & 6: Inhalation (6 seconds): Exhalation ratio (6 seconds)
Rest/gap has been given between each pranayama

**TABLE 3:**
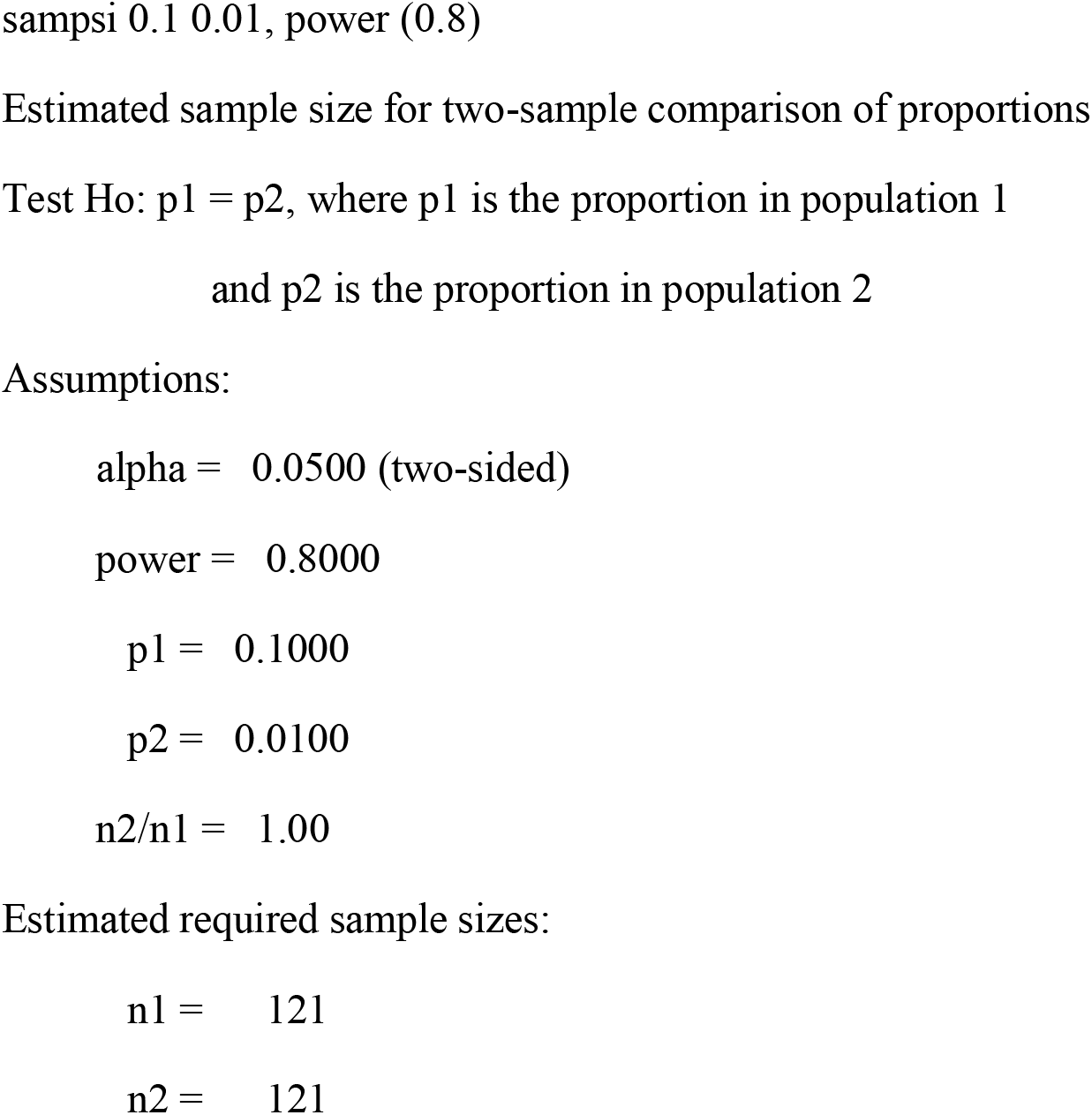
Sample Size Calculation in Stata

### Modules

In Yoga tradition, Pranayama includes not only deep breathing but also rhythmic, controlled respiration with awareness, which has four phases: inhalation, retention inside, exhalation and retention outside. The duration of the four phases of Pranayama was laid out based on traditional Yogic texts. The specially designed Pranayama modules included preparatory Yoga practices for 9 minutes, Pranayama practice for 16 minutes and meditation for 5 minutes in the morning session, and 15 minutes of breathing exercises in the evening session. Detailed procedure to perform the protocol has been provided as supplementary data (Annexure-II)

These modules were also publicly hosted (https://www.youtube.com/watch?v=RD1huWS_7w8&feature=youtu.be). The intervention group was provided this link from the beginning for their better understanding and aid in practice.

### Statistical Analysis

Data collected was entered on excel and validated. In bivariate analysis, categorical variables were presented as frequencies (%), continuous data as mean (with Standard Deviation) or median. Mean difference in pre-post intervention was examined for statistical significance using Fisher’s exact test. A P-value of 0.01 was considered significant. Data was analyzed using IBM Statistical Package for the Social Sciences (SPSS) Version 20.0. (Armonk, NY: IBM Corp).

## RESULT

### Socio demographic characteristics

Frontline Health Care Professionals were randomly categorized into Intervention and Control groups, comprising 123 and 127 participants respectively. Both groups had comparable demographic characteristics, with age ranging from 19 years to 65 years (Table4). The proportion of participants with co-morbid conditions was comparable in both the groups (Table 4).

**TABLE 4:** Socio Demographic Profile Of 250 HCPs

| **Variables** | **Experimental Grp**<br>**N (%)** | **Control Grp**<br>**N (%)** |
| --- | --- | --- |
| **TOTAL, N=250** | 123 (100.0%) | 127 (100.0%) |
| **Age, Years, Mean± SD** | 34.9 + 10.2 | 33.7 ±10.4 |
| 18-35 | 74 (60.16) | 70 (55.12) |
| 36-50 | 40 (32.52) | 49 (38.58) |
| 51-65 | 9 (7.32) | 8 (6.30) |
| **Gender, N (%)** |  |  |
| Male | 67 (54.47) | 65 (51.18) |
| Female | 56 (45.53) | 62 (48.82) |
| **Diet, N (%)** |  |  |
| Veg. | 69 (56.10) | 52 (40.94) |
| Mixed diet | 54 (43.90) | 75 (59.06) |
| **Exposure to COVID-19, N (%)** |  |  |
| In-directly Exposed | 86 (69.92) | 87 (68.50) |
| Directly Exposed | 37 (30.01) | 40 (31.50) |
| **Health Status, N (%)** |  |  |
| HCPs (Healthy) | 102 (83) | 106 (84) |
| HCPs (with Co-morbidities) | 21 (17) | 21 (16) |
| **Note: Grp: Group; N: Number; Veg.: Vegetarian** |  |  |

### Pranayama-Intervention Response on Perceived Stress Scores

The PSS Scores of the Intervention and Control groups at baseline were comparable at 13.84 + 6.62 (intervention), and 14.11 + 7.13 (Control) (Table 5).The preand postintervention average PSS score of the Interventional group (n =123) was 13.84 + 6.62 and 10.67 + 7.24 respectively. The pre- and post-intervention average PSS score of the Control group, (n =127) was 14.11 + 7.13 and 13.40 + 8.66 respectively. The mean difference of- 2.46 (CI: -4.6 to -0.26) between the two groups (intervention and control) is highly significant (P-value: 0.028) implying a reduction in Perceived stress in intervention group as compared to controls.

**TABLE 5:** Tabular representation describes Pre and Post Intervention of PSS, WHO Well Being, WHO QOL Score with Physical, Psychological, Social and Environmental domain in Interventional group compare to Control.

|  | No. of<br>Subjects | Pre-Intervention | Post-Intervention | Diff.<br>(Post Pre) | Mean Diff.<br>(Int. Cntrl),CI | P -Value |
| --- | --- | --- | --- | --- | --- | --- |
| PERCEIVED STRESS SCORE |  |  |  |  |  |  |
| INTERVENTIONAL | 123 | 13.84 ± 6.62 | 10.67 ± 7.24 | -3.17 ± 8.7 | -2.46 | 0.028** |
| CONTROL | 127 | 14.11 ± 7.13 | 13.40 ± 8.66 | -0.71 ± 8.9 | (-4.6 – -0.26) |  |
| WHO WELL BEING INDEX (MENTAL HEALTH SCALE) |  |  |  |  |  |  |
| INTERVENTIONAL | 123 | 18.29 ± 5.94 | 18.31 ± 7.51 | 0.016 ± 1.7 | -0.24 | 0.79 |
| CONTROL | 127 | 19.09 ± 4.77 | 19.35 ± 5.64 | 0.26 ± 1.3 | (-1.58 – 2.06) |  |
| WHO QUALITY OF LIFE SCORE |  |  |  |  |  |  |
| INTERVENTIONAL | 123 | 93.26 ± 15.27 | 96.34 ± 15.10 | 3.08 ± 3.78 | 2.78 | 0.17 |
| CONTROL | 127 | 87.31 ± 16.91 | 87.61 ± 17.27 | 0.29 ± 4.2 | (-6.8 – 1.3) |  |
| WHO QUALITY OF LIFE (PHYSICAL DOMAIN SCORE) |  |  |  |  |  |  |
| INTERVENTIONAL | 123 | 23.70 ± 4.57 | 23.94 ± 3.72 | 0.24 ± 1.04 | 0.64 | 0.29 |
| CONTROL | 127 | 22.51 ± 4.94 | 22.11 ± 4.44 | -0.4 ± 1.17 | (-1.8 0.5) |  |
| WHO QUALITY OF LIFE (PSYCHOLOGICAL DOMAIN SCORE) |  |  |  |  |  |  |
| INTERVENTIONAL | 123 | 22.02 ± 4.83 | 22.78 ± 5.06 | 0.77 ± 1.24 | 1.52 | 0.019** |
| CONTROL | 127 | 20.35 ± 4.44 | 19.60 ± 4.87 | -0.75 ± 1.17 | (-2.8 – -0.25) |  |
| WHO QUALITY OF LIFE (SOCIAL DOMAIN SCORE) |  |  |  |  |  |  |
| INTERVENTIONAL | 123 | 10.2 ± 2.46 | 11.20 ± 2.9 | 0.9 ± 0.63 | -0.12 | 0.76 |
| CONTROL | 127 | 9.57 ± 2.4 | 10.69 ± 3.06 | 1.12 ± 0.71 | (-0.69 – 0.94) |  |
| WHO QUALITY OF LIFE (ENVIRONMENTAL DOMAIN SCORE) |  |  |  |  |  |  |
| INTERVENTIONAL | 123 | 37.33 ± 7.03 | 38.41 ± 5.98 | 1.09 ± 1.62 | 0.68 | 0.48 |
| CONTROL | 127 | 34.81 ± 7.61 | 35.21 ± 7.47 | 0.4 ± 0.87 | (-2.64 – 1.26) |  |
| Diff. : Difference; CI: Confidence Interval, * indicate significance |  |  |  |  |  |  |

### Pranayama-Intervention Response on WHO Well-Being Index

The WHO Well-Being Index scores of study participants at baseline were 18.29 + 5.94(intervention), and 19.09+ 4.77 (Control) and thus comparable (Table 5). The preintervention average WHO Well-being index score of the intervention group (18.29 + 5.94) increased to 18.31 + 7.51 at end-line. The pre-intervention average WHO Well-being index score of the control group (19.09 + 4.77) increased at post-intervention to 19.35 + 5.64. The wellbeing index in Interventional group intervention showed a marginal increase.

### Pranayama-Intervention Response on Quality of Life (WHOQOL)

The WHOQOL scores of study participants at baseline were 93.26+ 15.27 (intervention), and 87.31 + 16.91 (Control) and thus comparable (Table 5).

The average WHO **Quality of Life score (**WHOQOL score) of the study participants (Interventional, number =123) increased from 93.26+ 15.27 to 96.34 + 15.10 post-intervention. In contrast, the change in Control group was marginal, from 87.31 + 16.91 to 87.61 + 17.27 (Table 5). This difference is not statistically significant at our sample size, however the overall quality of life showed improvement in the intervention group.

A domain wise analysis (Psychological, Social and Environmental) of scores on quality of life of participants was done. The results are different for all four domains

- **Physical Domain of WHO QOL**

The scores of study participants on Physical questionnaire scale at baseline were 23.70 + 4.57 (intervention), and 23.94 + 3.72 (Control) and thus comparable (Table 5). In the intervention group, the pre-intervention average WHOQOL physical score was 23.70 + 4.57, which increased post-intervention to an average of 23.94 + 3.72; in contrast, in the control group, preintervention average WHOQOL score was 22.51 + 4.94 declined somewhat post-intervention to 22.11 + 4.44. The increase in WHOQOL physical domain score in the intervention group was not significant.

- **Psychological Domain of WHO QOL**

In the intervention group, the average WHOQOL psychological score was 22.02 + 4.83 at baseline which increased to an average of 22.78 + 5.06 post-intervention. In contrast, controls had a pre-intervention average WHOQOL score of 20.35 + 4.44 which declined somewhat to 19.60 + 4.87 post-intervention. The increase in WHOQOL psychological score in the intervention group over the control (Mean diff: -1.52) was significant (P-value: 0.019)

- **Social Domain of WHO QOL**

In the intervention group, the average WHOQOL social score was 10.20 + 2.46 at baseline which increased to an average of 11.20 + 2.9 post-intervention. Controls had a pre-intervention average WHOQOL score of 9.57 + 2.4 which increased somewhat to 10.69 + 3.06 post-intervention. The WHO-QOL Social domain score showed a less increase in the Intervention group, while it increased more in the Control group at post-intervention. However, these changes were not significant (p-value: 0.76) and overall mean difference was negative.

- **Environmental domain**

In the intervention group, the average WHOQOL environmental score was 37.33+ 7.03 at baseline which increased to an average of 38.41+ 5.98 post-intervention (Table 5). In contrast, controls had a pre-intervention average WHOQOL score of 34.81 + 7.61 which increased somewhat to 35.21 + 7.47 postintervention. The WHO-QOL Environmental score showed an increase in intervention group. The difference was however, not statistically significant at the sample strength (p-value: 0.48).

## DISCUSSION

Our study revealed that twice daily practice of Pranayama session for 28 days among Healthcare Professionals had beneficial effects as shown by ssignificantly reduced Perceived stress score; increase in overall WHO-Quality of Life Score and significant increase in psychological domains of WHOQOL score, suggesting a positive effect of Pranayama sessions on HCPs on mental health. Individual scores on PSS can range from 0-40 with higher scores indicating higher perceived stress. Similarly, high scores consider the higher wellness and better quality of life.

The WHO Quality-of-life score also showed increase in the intervention group as compared to the controls (mean difference 2.78, p-value: 0.17) but not significantly. The WHO QOL is the index of quality of life with 4 domains: Physical, psychological, social and environmental aspects of quality of life of an individual. Each domain considers a different aspect of quality of life of a person. The physical considers the physical activity related quality of life. Then psychological domain covers the mental health aspect. Social and environmental domains assess the social wellness and environmental positivity of a person.

The study in Italy on 595 HCPs with Perceived stress scale scores were, males (mean = 15.38; SD = 6.65) and females (mean = 19.56; SD = 7.06) (Babore A et.al.2020) comparable to current study. Various studies conducted on HCPs in different countries like India, China, Turkey etc to understand Quality of Life (WHO-QOL) and stress levels (PSS) agree with the results of present study (Wilson W et.al.2020; Tian T et.al.2022; Korkmaz S et.al.2020) To our knowledge, this is among the very few studies on the effect of pranayama in reduce stress; improve well-being and quality of life among frontline HCPs during pandemic. Our results are consistent with other comparable studies.

There are numerous explanatory pathways for the improvement with Pranayama on stress. Stress is known to suppress immune function and increase susceptibility to infections (Dhabhar FS et al.2009). Chronic stress is associated with global immuno-suppression. Increasing duration of stress can result in a shift from potentially adaptive changes to potentially detrimental changes, initially in cellular immunity and later and more broadly in immune function (Segerstrom SC et.al2004^)^. Pranayama practices have been found to have a direct impact on vagus Nerve stimulation (Howland RH,2014), Vagus nerve Stimulation has been found to have a direct effect in managing stress and diseases born out of it (Howland RH,2014). The efficacy of our especially designed Pranayama protocol could be because of the above-mentioned molecular mechanisms, whose modalities of action need further exploration. This study in a bundled analysis has also revealed that COVID-19 infection in the intervention group tends to be mild and asymptomatic, thus strengthening the conclusion of a positive effect of Pranayama in preventing COVID 19 infection (Sarwal et al. 2022).

## STRENGTHS AND LIMITATIONS

The strengths of the present cross-sectional study are: it was conducted in 5 different hospitals of Delhi (dedicated for COVID-19 patients) with a reasonable (250) sample size. There are, however, some limitations pertaining to the study. Due to pandemic conditions, video-conferencing was used to administer Pranayama protocols. Prolonged working hours did lead to lowered interest in HCPs for Pranayama classes. PSS, WHO Wellbeing Index, WHO-QOL has been assessed only through a self-reported questionnaire. A multi-centric study with a large sample size, in-person instruction and a longer duration may give answers with a greater validity on the pranayama protocol. More studies are needed to extend and verify the generalization of the present results.

## CONCLUSION

The simple intervention of Pranayama may be effective in lowering stress, improving mental health and quality of life in frontline Healthcare Professionals (HCPs). This protocol is open source, and can be self-governed or supervised from a distance with minimal cost. Therefore, this study emphasizes on the need for practice of the Pranayama modules by those routinely exposed to psychological distress, such as frontline Hospital’s staff, care givers, and general public. Front line HCPs should take to the practice of Pranayama to improve their mental health, and reduce stress. Funding Agency: Morarji Desai National Institute of Yoga, Ministry of AYUSH, Government of India

**FIGURE 1:**
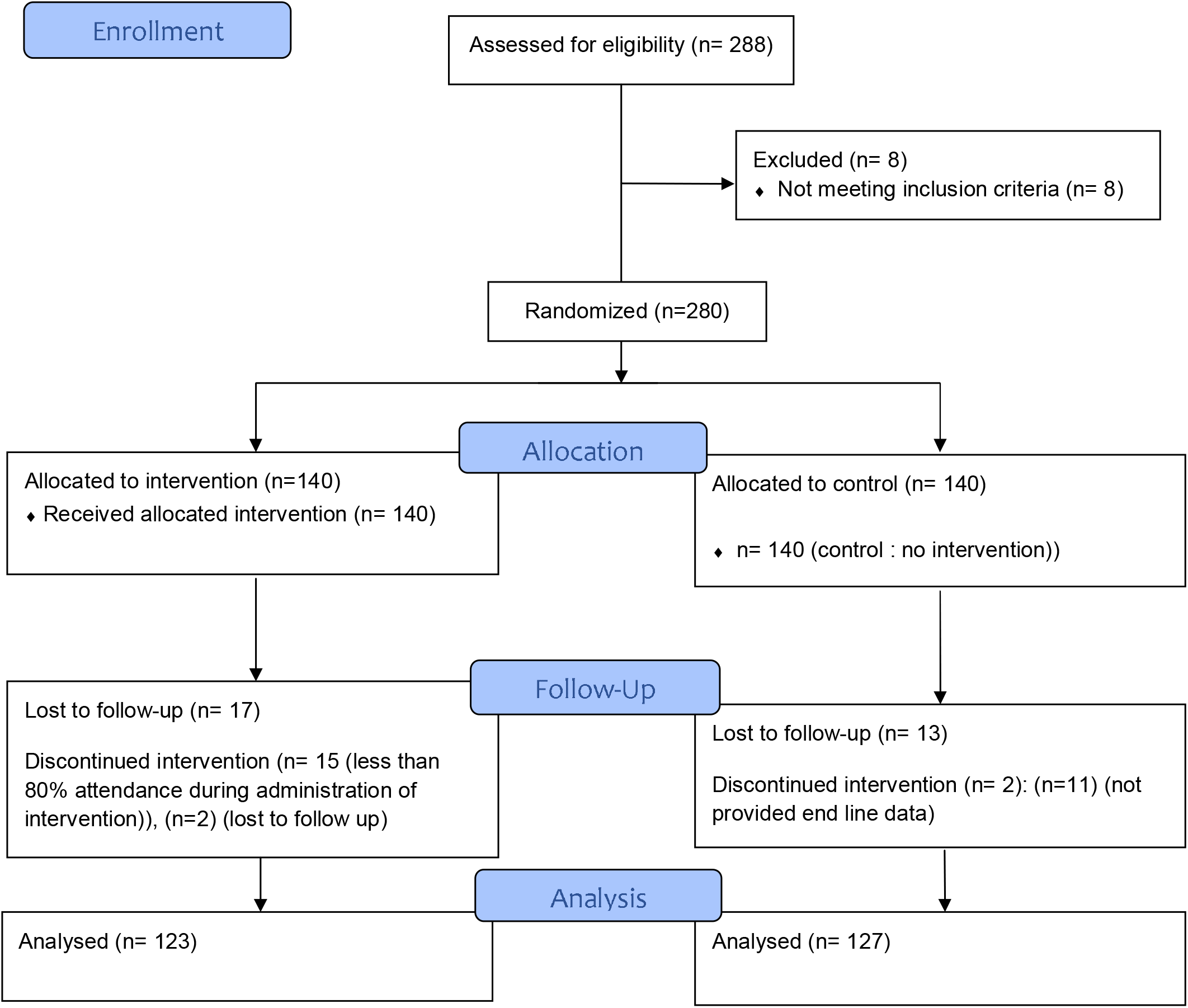
Flow Diagram.

## Supporting information

Annexure II

## Data Availability

All data produced in the present study are available upon reasonable request to the authors

https://www.youtube.com/watch?v=RD1huWS_7w8&feature=youtu.be

## ACKNOWLEDGEMENTS

The authors would like to thank Vaidya Rajesh Kotecha, Secretary, Ministry of AYUSH and Chairman, Governing Council, MDNIY for his support and guidance. We would also like to extend our thanks to Shri P.N. Ranjit Kumar, Joint Secretary, AYUSH and Chairperson, Scientific Advisory Committee (SAC), MDNIY and all other members of SAC for their valuable inputs and encouragement. We also thank Dr. Shivam Pandey, Scientist-1, Dept. of Biostatistics, AIIMS for his help in doing statistical analysis of the research work. We would also like to offer our special thanks to Dr. R.K. Manchanda, Director, Directorate of AYUSH Govt. of NCT Delhi for his support in arranging the much-needed participants for the study and Dr. I.N. Acharya, Program Officer (YT) of MDNIY for helping supervise the administration of the modules. The authors also thank medical superintendents, doctors and staff of all 5 hospitals for their active cooperation. We would like to thank 5 yoga instructors (Mr. Sahil, Mr. Sandeep, Ms. Aishwarya, Ms. Sunita and Ms. Kranti) and Data entry operator (Ms. Nishu) for administration of yoga modules and entry of data respectively.

## Supporting information

- The study was conducted in the following hospitals:
- Dr. Hedgewar Arogya Sansthan, New Delhi
- Guru Tegh Bahadur Hospital, Dilshad Garden, Delhi
- Deep Chand Bandhu Hospital, Delhi
- Govt. of NCT Delhi Dispensary, Batla House, Jamia Nagar, New Delhi
- Sultanpuri Quarantine Centre, Sultanpuri, New Delhi.

## ANNEXURES

Annexure I: Psychological Questionnaires

Annexure II: Detailed procedure of Pranayama Module

## ANNEXURE-1

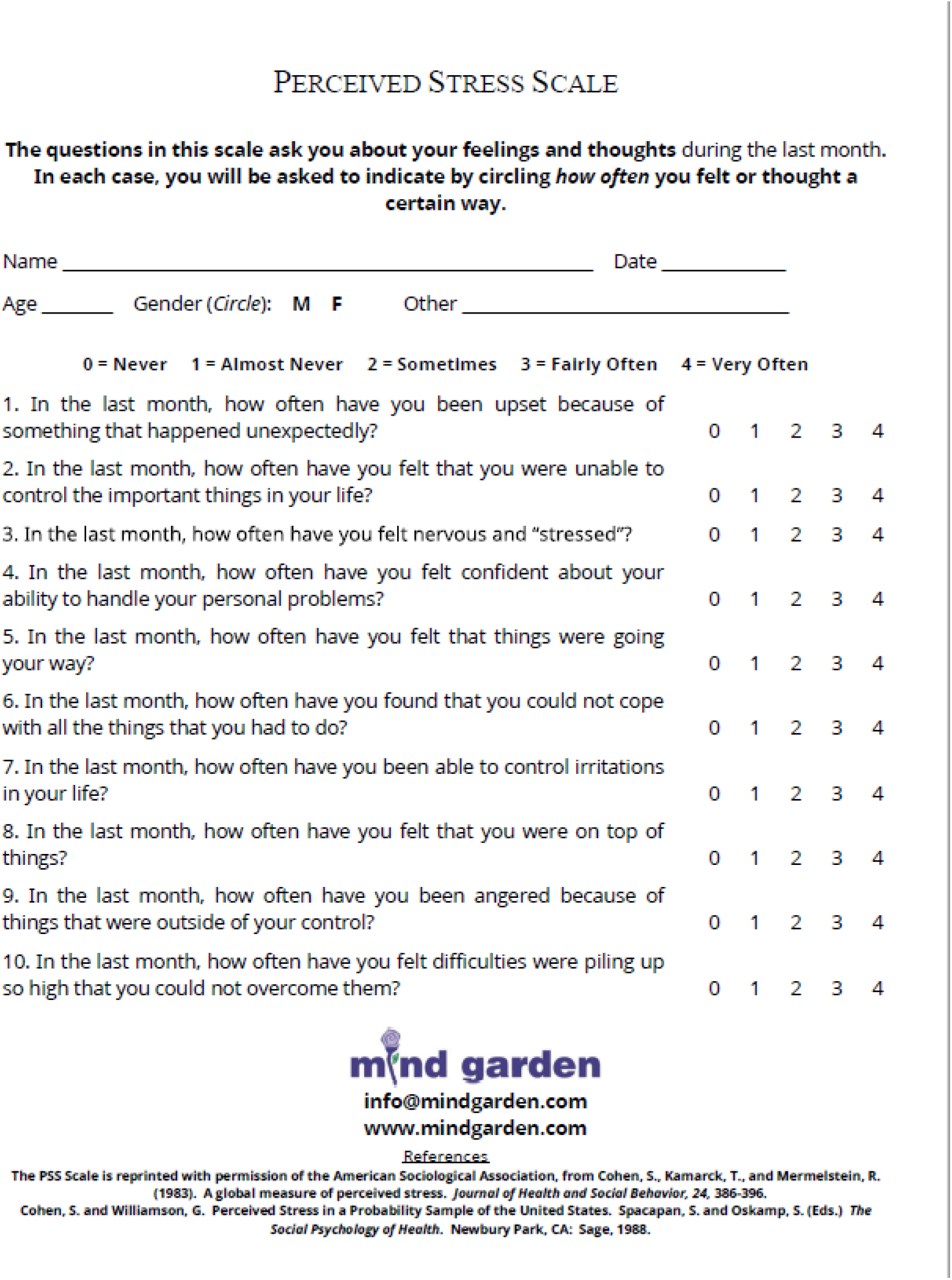

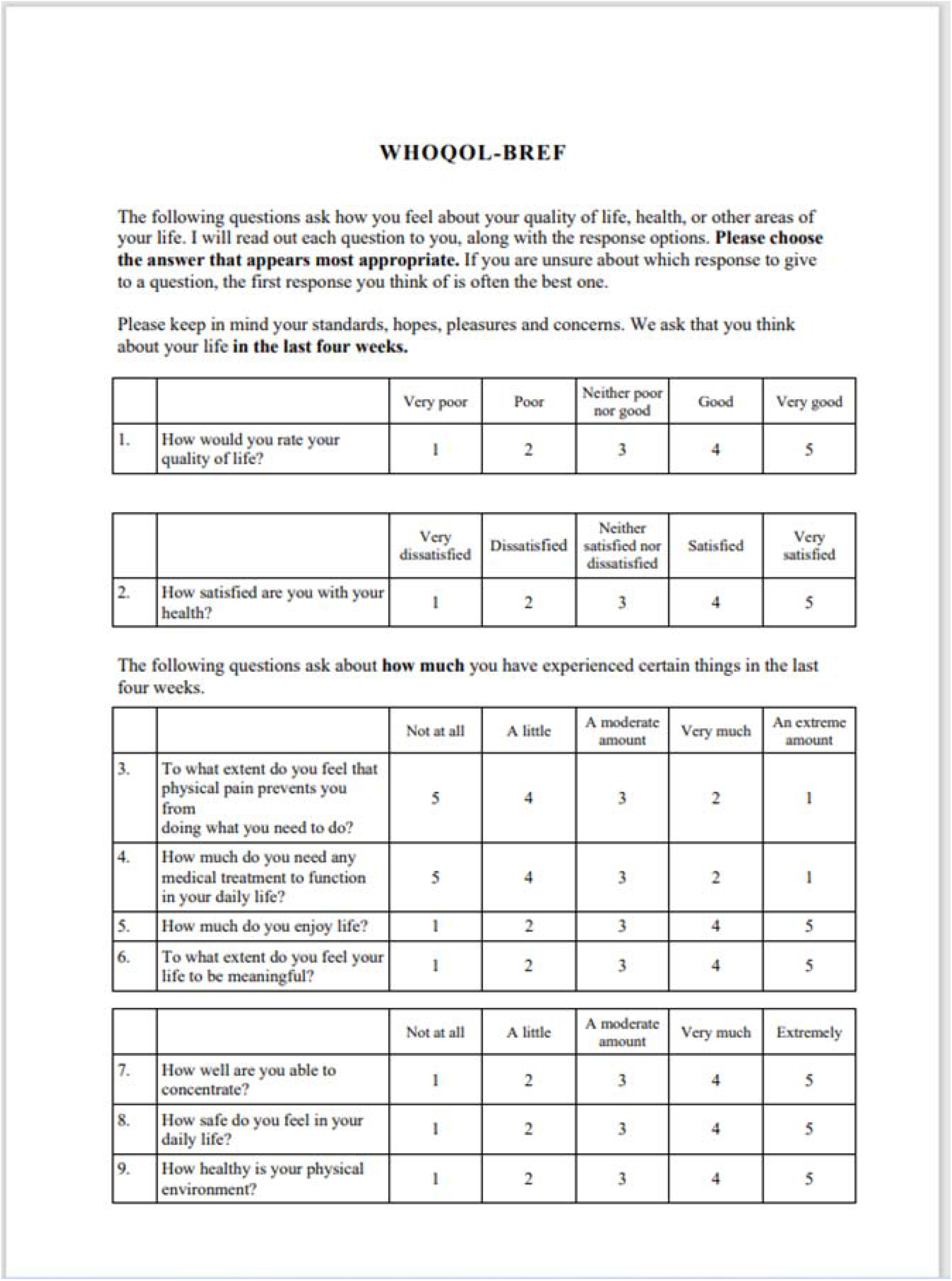

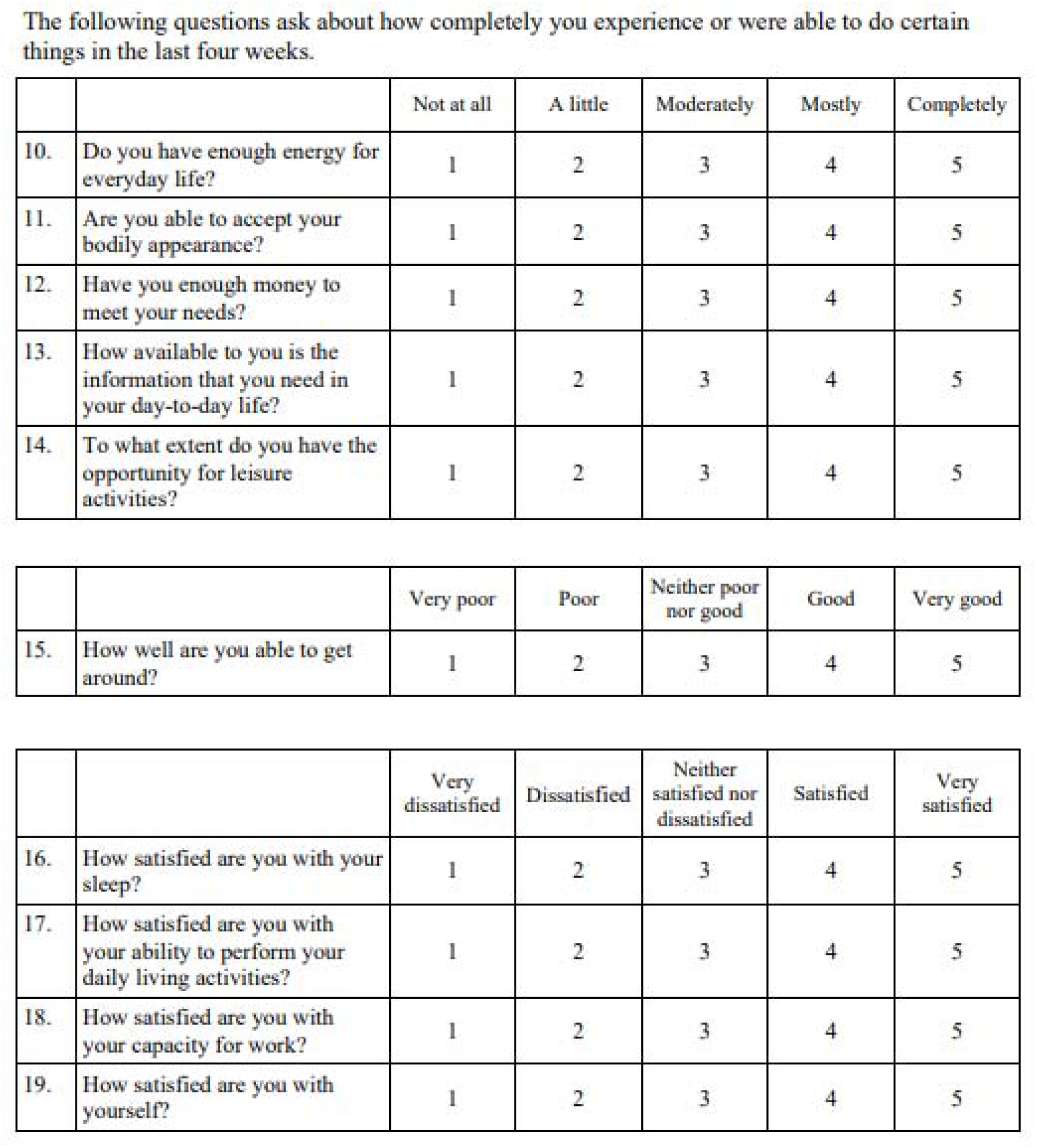

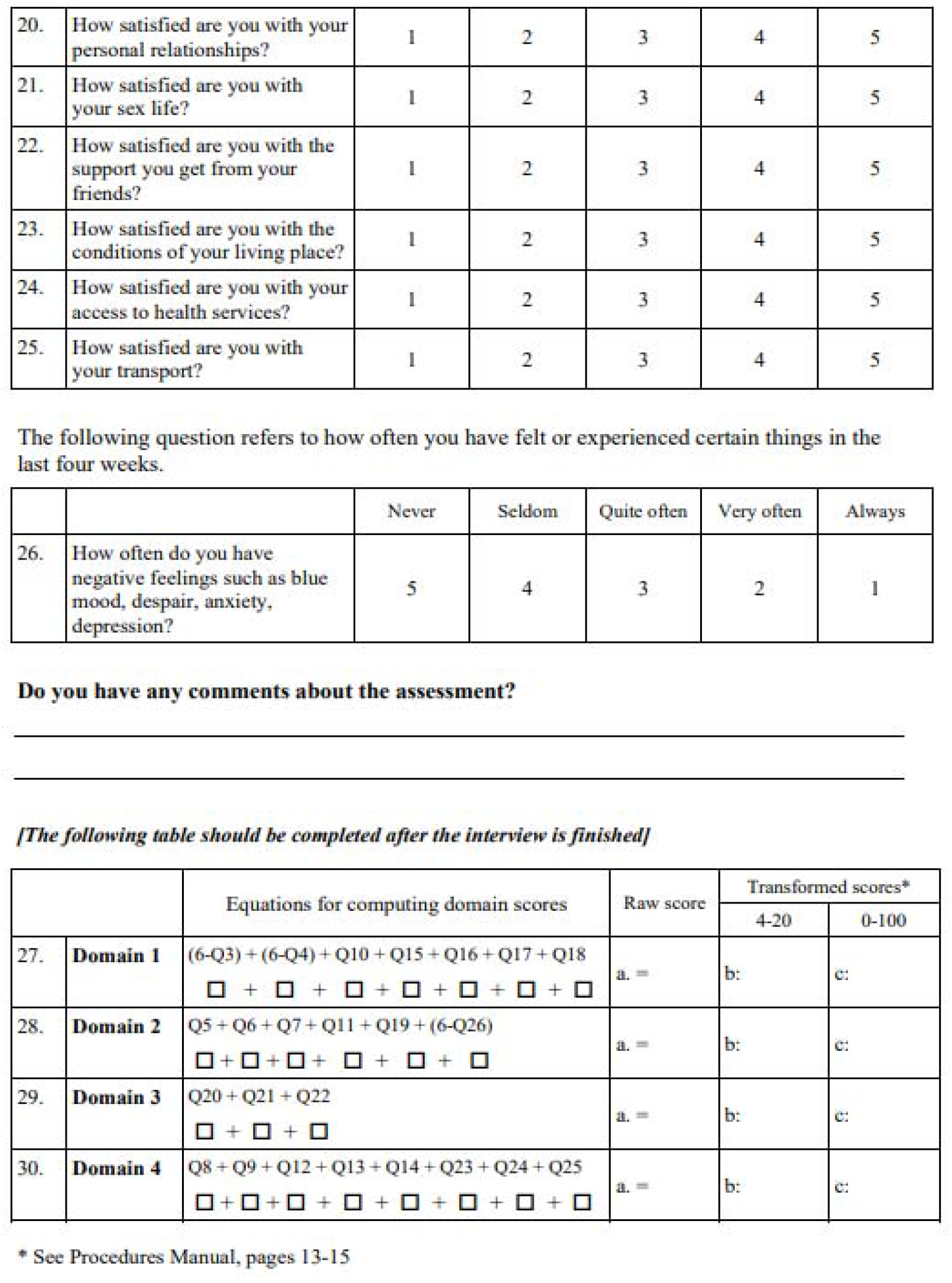

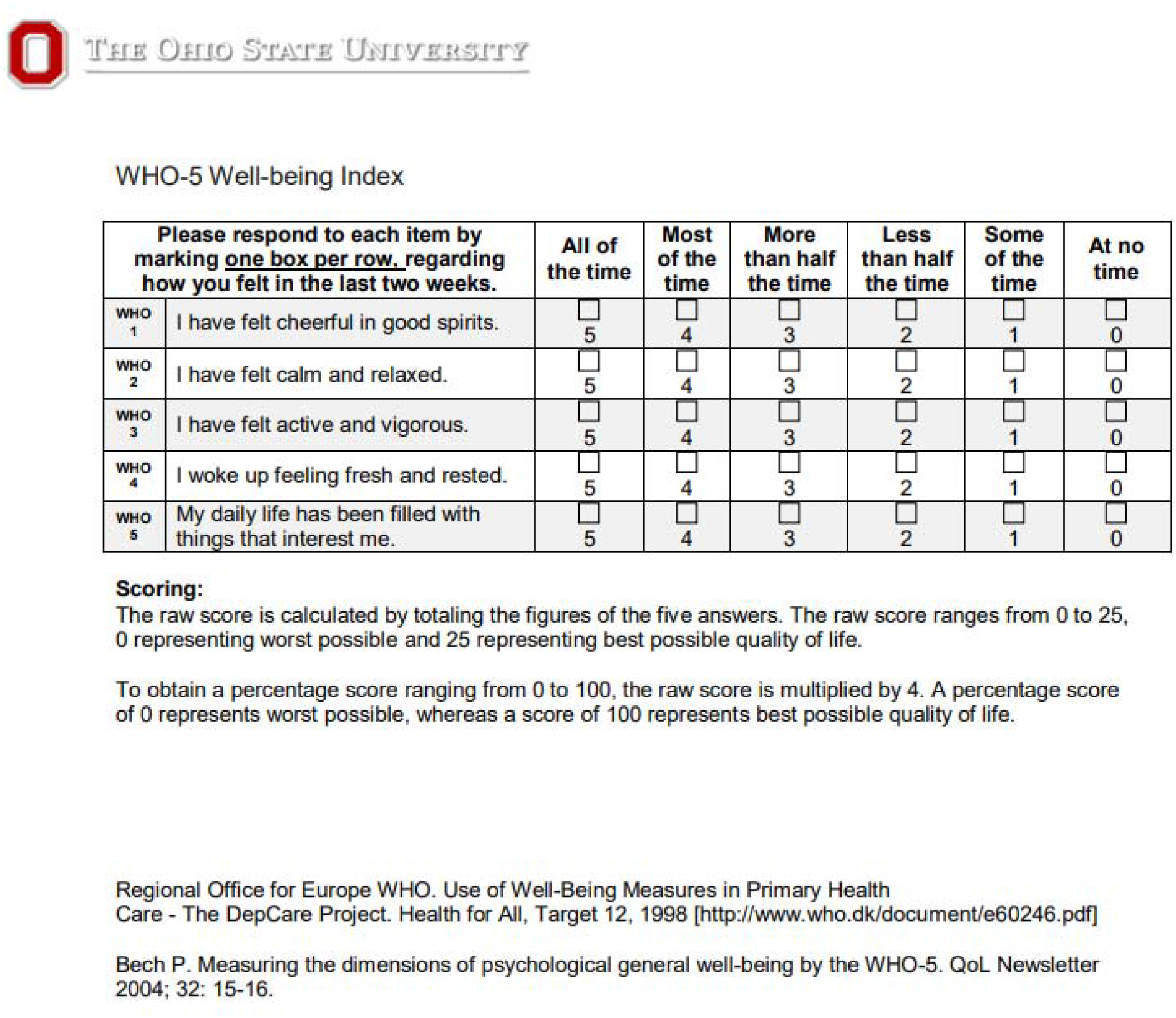

