## Supplementary material for "Effect of Pranayama on Perceived Stress, Well Being and Quality of Life of Frontline Healthcare Professionals on Covid-19 Duty: A Quasi-Randomized Clinical Trial": Annexure II

**ANNEXURE-1: DETAILED PRANAYAMA PROTOCOL**

***GUIDELINES FOR PROTOCOL:***

BEFORE THE PRACTICE:

- Śauca (SHAUCH) means cleanliness - an important prerequisite for Yoga practice. It includes cleanliness of surroundings, body and mind.
- Yoga practice should be performed in a calm and quiet atmosphere with a relaxed body and mind.
- Yoga practice should be done on an empty stomach or light stomach. Consume small amount of honey in lukewarm water if you feel weak.
- Bladder and bowels should be empty before starting Yogic practices.
- It is advisable to use a Yoga mat, mattress, durrie or folded blanket for the practices.
- Light, comfortable, not too tight and preferably cotton clothes to facilitate easy movement of the body.

DURING THE PRACTICE:

- Practice should be done in comfortable seated posture such as Sukhasana, Ardha-padmasana, Padmasana, Vajrasana or can be done sitting on the chair.
- Practice sessions should start with a prayer or an invocation as it creates a conducive environment to relax the mind.
- Yogic practices shall be performed slowly, in a relaxed manner, with awareness of the body and breath.
- Breathing should always be through the nostrils unless instructed otherwise.
- Breathe should be deep, slow and rhythmic.
- Adopt proper mudra during the practice as shown in the video. Like Jnana Mudra, Pranayama Mudra, Dhyana Mudra.
- Perform the practices according to one’s capacity. In case of difficulty in practicing recommended 6 seconds, one can start practicing for 4 seconds.
- It takes time to get good results, therefore persistent and regular practice is very essential.
- Yoga session should end with meditation/ deep silence etc.

AFTER PRACTICE:

- Bath may be taken only after 20-30 minutes of Yoga practice.
- Food may be consumed only after 20-30 minutes of Yoga practice.

**MOR NING SESSION**

Prayer:

Sit in a comfortable seated posture, keeping back and neck straight.

Close your eyes. Join palms in Namaskar Mudra.

Let’s chant Om three times

*O……M…….*

*O…...M…….*

*O…...M…….*

Shantih… Shantih… Shantih…

Gently open your eyes and take your hands back.

**VATANETI**

- Close your eyes. Place palms in Jnana Mudra on the respective knees.
- Slightly chin upwards.
- Inhale deeply, then start rapid inhalation and exhalation for 30 seconds.
- Deep exhale and relax…. (Feel the change in the nasal passage and forehead)
- Start Second round ….
- Deep exhale and relax…..
- Start Third round…..
- Deep exhale and relax…..

**KAPALABHATI**

- Keep the back and the neck straight.
- Inhale deeply and expel the breath with forceful contractions of the abdominal muscles.
- Active exhalation and passive inhalation for 30 seconds.
- Start the First Round
- Now exhale deeply and relax (feel the peace).
- Start the second round of Kapalbhati.
- Exhale deeply and relax.
- Now, start the third and the final round.
- Exhale deeply and relax.

**SHAVASANA**

- Lie down on the back.
- Spread the legs and hands apart.
- Palms facing upwards.
- Close the eyes
- Relax the whole body.
- Observe the normal breathing.
- After one minute
- Now join the legs, stretch your arms upwards
- Stretch the whole body.

**DEEP BREATHING**

- Inhale deeply through nostrils with the expansion of abdominal, thoracic and clavicular region.
- While exhaling, relax abdominal, thoracic and the clavicular region in a reverse manner.
- This is one round / This is round one.
- Repeat for nine more rounds.
- Inhale 1,2,3,4,5,6
- Exhale 1,2,3,4,5,6,
- Completely exhale and relax the body and mind.
- Join your legs turn by the side and come back to the sitting position.

**NADI SHODHANA**

- Sit in any comfortable posture.
- Close the eyes.
- Keep the back and the neck straight.
- Adopt Jnāna mudra.
- Now, Adopt Nāsāgra or Pranayama mudra with the right hand.
- Exhale from both the nostrils.
- Now, close the right nostril and inhale through the left for 6 seconds.
- Close both nostrils retain for 3 seconds and exhale through the right for 6 seconds.
- After exhalation again close both nostrils and retain for 3 seconds.
- Now, inhale through the right for seconds, retain for 3 seconds and exhale through the left.
- After exhalation retain for 3 seconds.
- This is one round.
- Repeat for nine more rounds.
- Purak	Kumbak
- Rechak	Shunyk
- Inhale 1, 2, 3, 4, 5, 6
- Retain 1,2,3
- Exhale 1, 2, 3, 4, 5, 6,
- Retain 1,2,3
- Inhale 1, 2, 3, 4, 5, 6
- Retain 1,2,3
- Exhale 1, 2, 3, 4, 5, 6
- Retain 1,2,3
- Inhale from left 2, 3, 4, 5, 6
- Retain 1,2,3
- Exhale from right 2, 3, 4, 5, 6,
- Retain 1,2,3
- Inhale from right 2, 3, 4, 5, 6
- Retain 1,2,3
- Exhale from left 2, 3, 4, 5, 6
- Retain 1,2,3

While exhaling through the left nostril bring the right hand back.

**UJJAYI PRANAYAMA**

- Keep the back and the neck straight.
- Place your hands on the knees in Jnāna mudra.
- While slightly contraction of the throat, Inhale deeply through the nose while making a hissing sound for 6 seconds.
- Retain the breath closing both the nostrils for 3 seconds.
- Deep exhale through both the nostrils for 6 seconds.
- Again retain for 3 seconds.
- This is one round.
- Repeat for nine more rounds.
- Inhale 1, 2, 3, 4, 5, 6
- Retain 1,2,3
- Exhale 1, 2, 3, 4, 5, 6
- Retain 1,2,3
- Complete exhale and relax the body and mind.

**BHRAMARI PRANAYAMA**

- Keep the eyes closed and the hands in jnana mudra.
- Inhale deeply for 6 seconds and while exhaling make a deep, steady humming sound like a bee…for 9 seconds
- Start first round. Inhale 1,2,3,4,5,6
- Hmm……….
- This is one round
- Repeat for nine more rounds.
- Inhale 1, 2, 3, 4, 5, 6
- Hmmmmm….. ( till 9 seconds)

**DHYANA….**

- Keep your eyes closed.
- Deep breathe and exhale.
- Adopt dhyana mudra.
- Keep your spine erect.
- Keep the shoulder down and relax the elbows.
- Relax the whole body.
- Deep breath for 3 times.
- Meditate.

Slowly come out of the Dhyana.

Now join the palms together in namaskar mudra and brings your hands back.

Open your eyes.

Relax.

**EVENING SESSION:**

**SAVASANA (Corpse pose) – Pristhbhumi Tadasana**

- Lie down on the back.
- While inhaling, stretch the arms over the head.
- Stretch the whole body.
- Maintain the position with normal breathing.
- While exhaling, relax the whole body with proper gap between legs and hands, palms facing upwards.
- Relax the body for 3 breaths.
- Turn by the side and come back to the sitting position.

Prayer:

Sit in a comfortable seated posture, keeping back and neck straight.

Close your eyes. Join palms in Namaskar Mudra.

Let’s chant Om three times

*O……M…….*

*O…...M…….*

*O…...M…….*

Shantih… Shantih… Shantih…

Gently open your eyes and take/bring your hands back.

**ABDOMINAL BREATHING:**

- Keep back and neck straight.
- Place both the palms in Jnana mudra on respective knees.
- Now place right hand on the abdomen.
- Complete exhale through the nostrils.
- Inhale for 6 seconds, abdomen will move outward.
- Exhale for 6 seconds, abdomen will move inward.
- Keep awareness on the abdomen while inhaling and exhaling.
- Repeat for 14 more times.
- Inhale 1,2,3,4,5,6
- Exhale 1,2,3,4,5,6

Relax and bring the right hand back.

**THORACIC BREATHING (Chest Breathing):**

- Keep the spine erect.
- Place both palms on the chest.
- Observe the breath concentrate on the thoracic region.
- Inhale for 6 seconds, feel expansion of thoracic region.
- Exhale for 6 seconds, relax the thoracic region.
- Repeat for 14 more times / Repeat for 9 more times.
- Inhale 1,2,3,4,5,6
- Exhale 1,2,3,4,5,6

Relax and bring the hands back.

**CLAVICULAR BREATHING: (shoulder breathing)**

- Place both the palms on the opposite shoulder blades.
- Inhale for 6 seconds and expand the upper region of lungs around the collar bone.
- Exhale slowly for 6 seconds; relax the neck region.
- Repeat for 14 more times.
- Inhale 1,2,3,4,5,6
- Exhale 1,2,3,4,5,6

Relax and bring the hands back.

**DEEP BREATHING: (In shavasana position)**

- Lie down in Shavasana and relax the whole body.
- Inhale slowly and deeply for 6 seconds, allowing the abdomen, thoracic and collar bone region to expand fully.
- Exhale deeply for 6 seconds and relax the abdomen, thoracic and collar bone region in the reverse way.
- Repeat for 14 more times.
- Inhale 1,2,3,4,5,6
- Exhale 1,2,3,4,5,6

**SHAVASANA WITH ABDOMINAL BREATHING:**

- Relax the whole body in shavasana with legs and hands comfortably apart.
- Keep the awareness on the abdominal region during inhalation and exhalation.
- Take deep and slow breath.
- Relax Completely ……..

R… e… l…a….x…

D…h…y….a…n….a….
